# Reasons for mosquito net non-use in malaria-endemic countries: A review of qualitative research published between 2011 and 2021

**DOI:** 10.1101/2023.05.16.23290037

**Authors:** Hadiza Isa Ladu, Umar Shuaibu, Justin Pulford

## Abstract

Mosquito nets, particularly insecticide-treated nets [ITNs], are the most recommended method of malaria control in endemic countries. However, many individuals do not use them as advised. The current paper expands on a previous review published in 2011 which highlighted a need for more qualitative research on the reasons for mosquito net non-use. We present a systematic review of qualitative research published in the past decade to assess the growth and quality of qualitative papers about net non-use and examine and update the current understanding.

A comprehensive literature search was carried out in MEDLINE, CINAHL, and Global Health, in addition to a citation search of the initial review. Relevant papers were screened and discussed. The critical appraisal assessment tool was used to ensure quality. Thematic synthesis was used to extract, synthesise, and analyse study findings.

Compared to the initial review, the results showed a ten-fold increase in qualitative research on the reasons for mosquito net non-use between 2011 and 2021. In addition, the quality of the research has improved, with more than 90% of the papers receiving high scores, using the critical appraisal assessment tool. The reported reasons for non-use were categorised into four themes Human factors, Net factors, Environmental/Lifestyle factors, and Administrative/Economic factors. More than two-thirds of the studies were carried out in Africa, with lead African researchers in African institutions.

Despite the distribution of free mosquito nets in malaria-endemic countries, new challenges to their use continue to emerge. The most common reasons for net non-use across all regions of Malaria endemic countries were discomfort and perceived ineffectiveness of nets. Technical challenges and improper net use dominated East and South African regions, signifying the need for dedicated and region-specific measures and strategies to ensure the continued usage of mosquito nets, particularly ITNs.

## INTRODUCTION

There were an estimated 247 million malaria cases across 84 malaria-endemic countries [MECs] in 2021, with more than 90% of the cases recorded in Sub-Saharan Africa. The mortality rate ranges between 0.3 – 2.2% and can be as high as 11-30% for severe malaria (1). Malaria infection may further affect an individual’s social and economic life. Many individuals in MEC are poor; thus, infection with the disease increases their financial burden through drug procurement and travel expenses to health centers or clinics. Other economic costs from the illness include loss of wages due to absence from work, and frequent infection can also impact the learning of children who miss school (2).

Malaria can be prevented and controlled in MEC through the routine use of mosquito nets (2, 3). There are two kinds of mosquito nets: insecticide-treated nets [ITNs], an “umbrella” term encompassing “all nets treated with an insecticide, insect growth regulator and or synergist” (4), and untreated mosquito nets. An untreated net protects the individual/s resting within against mosquito and other pest/insect bites (5, 6). While ITNs offer personal protection to those under the net and kill mosquitoes conferring a more comprehensive household/community benefit (7). An estimated 1.2 billion cases of malaria and 7.1 million deaths were averted between 2004 and 2019 in sub-Saharan Africa following antimalaria campaigns, with ITNs making the single most significant contribution accounting for an estimated 68% of these figures (8).

The cumulative shipments of all forms of mosquito nets to MEC increased from 436 million in 2010 to approximately 2.6 billion in 2021 (9). While access to ITNs may have improved in MEC over this time, this does not imply increased use, defined as “the proportion of the population that does sleep under a net” (8). For example, at least 72% of Sub-Saharan African households had at least one ITN in 2018; however, only 50% of the population at risk slept under an ITN the night before (10) despite WHO guidelines recommending continuous use of ITN nightly in Malaria endemic areas irrespective of climatic conditions (4). Nevertheless, ITN remains a cost-effective strategy for controlling and preventing malaria (11) and global support remains to increase ITN access and use in MEC, particularly in sub-Saharan Africa (2).

A previous review by Pulford et al. (12) examined reasons for net non-use as reported in the published literature. This review identified discomfort due to heat and perceived low density of mosquitoes as the primary reasons for net non-use (12), although Pulford et al. (12) concluded that more and better quality research examining reasons for ITN non-use was needed and especially qualitative studies. In the decade that has passed since the publication of this review, a wide range of research [inclusive of qualitative research] has been completed, and a more complex understanding of reasons for ITN non-use appears to be emerging. For instance, Perkins et al.(13) noted that people frequently imitate other people’s bed-net usage behaviour and Guglielmo et al.(14) draw on qualitative findings to challenge how ‘use’ and ‘compliance’ are even evaluated in the context of ITN use for malaria control. However, the more recent research effort’s scope, quality, and findings have yet to be reviewed.

In this paper, we present an up to date review of published qualitative research exploring mosquito net non-use conducted in the decade since the original review by Pulford et al. (12). Our specific objectives were to assess the extent and quality of qualitative research published between 2011-2021 and, drawing on the findings from this literature, update current understanding of reasons for mosquito net non-use in MEC. It is hoped the results may inform the design and selection of net-use promoting interventions.

## METHODOLOGY

We conducted a structured literature review of qualitative studies published between 2011 – 2021 which examined reasons for mosquito net non-use in MEC.

### Search strategy

A structured electronic search of peer-reviewed articles was conducted using the following three databases: MEDLINE, CINAHL, and Global health. An additional citation search of the original Pulford et al. review (12) was conducted to identify and retrieve relevant papers. The search terms used in the original Pulford et al. review (12) were adapted and applied as a baseline for the current study: [1] Malaria AND [2] Mosquito Bed Net AND [3] Non-use AND [4] Qualitative, with variants of the terms [Table1].

**Table 1:** Original reviews search terms, adjusted for year and study type.

| Search ID# | MeSH terms. |
| --- | --- |
| S8 | S7 AND S6 |
| S7 | “qualitative*” OR “focus*group” OR “interview*” OR “mixed method” |
| S6 | S4 AND S5 |
| S5 | “non-use*” OR “obstacle*” OR “misuse*” OR “disuse*” OR “challenge*” OR “neglect*” OR “abandon*” OR “barrier*” OR “use*” |
| S4 | S2 AND S3 |
| S3 | “Mosquito Bed Net*” OR “Mosquito Net*”<br>OR “Mosquito Bednet*” OR “Insecticid*treated” OR “ITN*” OR “LLIN*”<br>OR “Insecticide-treated Bednets” OR “Bednet*” |
| S2 | “Malaria*”[201-2021] |
| S1 | “Malaria*” |

### Study selection

All retrieved publications were imported into Endnote, and duplicates removed. Publications were initially screened by title and abstract against specified inclusion criteria, with the remaining publications subject to full-text review. All publications were independently reviewed for inclusion at both stages by two authors [HIL, US], with any disagreements resolved by discussion until consensus was reached. The inclusion criteria included studies: conducted in MEC, research focused on mosquito net non-use [as either primary or secondary objective], published in English between 2011 and 2021, employed qualitative methods or a mixed method study design with transparent reporting on qualitative methods and results.

### Quality appraisal

The Critical appraisal skill programme [CASP] checklist was utilised to assess the quality of the retrieved papers. CASP consists of ten structured questions that cover the following topics: study purpose, appropriateness of qualitative design, suitability of study design, sampling strategy, data collection method, ethical consideration, data analysis, findings, and study relevance [see Table 3]. Each set of questions has three scoring options: Yes (1), No (0), or Can’t tell (0). Each set of questions has hints to guide the researcher in evaluating papers. Studies with a cumulative score of eight and above were regarded as high quality, five to seven as medium quality, and below five as low-quality studies (15, 16). Two authors independently assessed all publications against the CASP criteria [HIL, US]. An agreement was reached between both authors on the CASP scoring for each paper.

### Data extraction and analysis

The lead researcher extracted the following information into an Excel file from each paper; Research title, authors [1^st^ and last], institution of lead author, year of publication, study objectives, study settings, country of focus, study participants, study type [qualitative alone or mixed study] and Information on data collection tools, analysis, and sampling methods. Finally, a summarised version of the key findings reported within each paper concerning reasons for ITN non-use was extracted.

Summarised versions of the key findings were analysed thematically. The first step was to become acquainted with the data. This entailed reading the extracted data repeatedly, identifying common themes pertinent to our study objectives, and highlighting related quotes that emphasise and elaborate on the themes. Subsequently, a coding framework was developed. The key findings data were then systematically coded against this framework and a final set of themes and associated sub-themes identified. All other extracted data were analysed using descriptive statistics as presented in the results section below.

## RESULTS

### Study selection

As depicted in Fig. 1(Attached file) the initial search yielded 892 articles, of which 81 met the inclusion criteria for a full-text review. Following full-text review, 39 articles were included in the final sample

**Figure 1:**
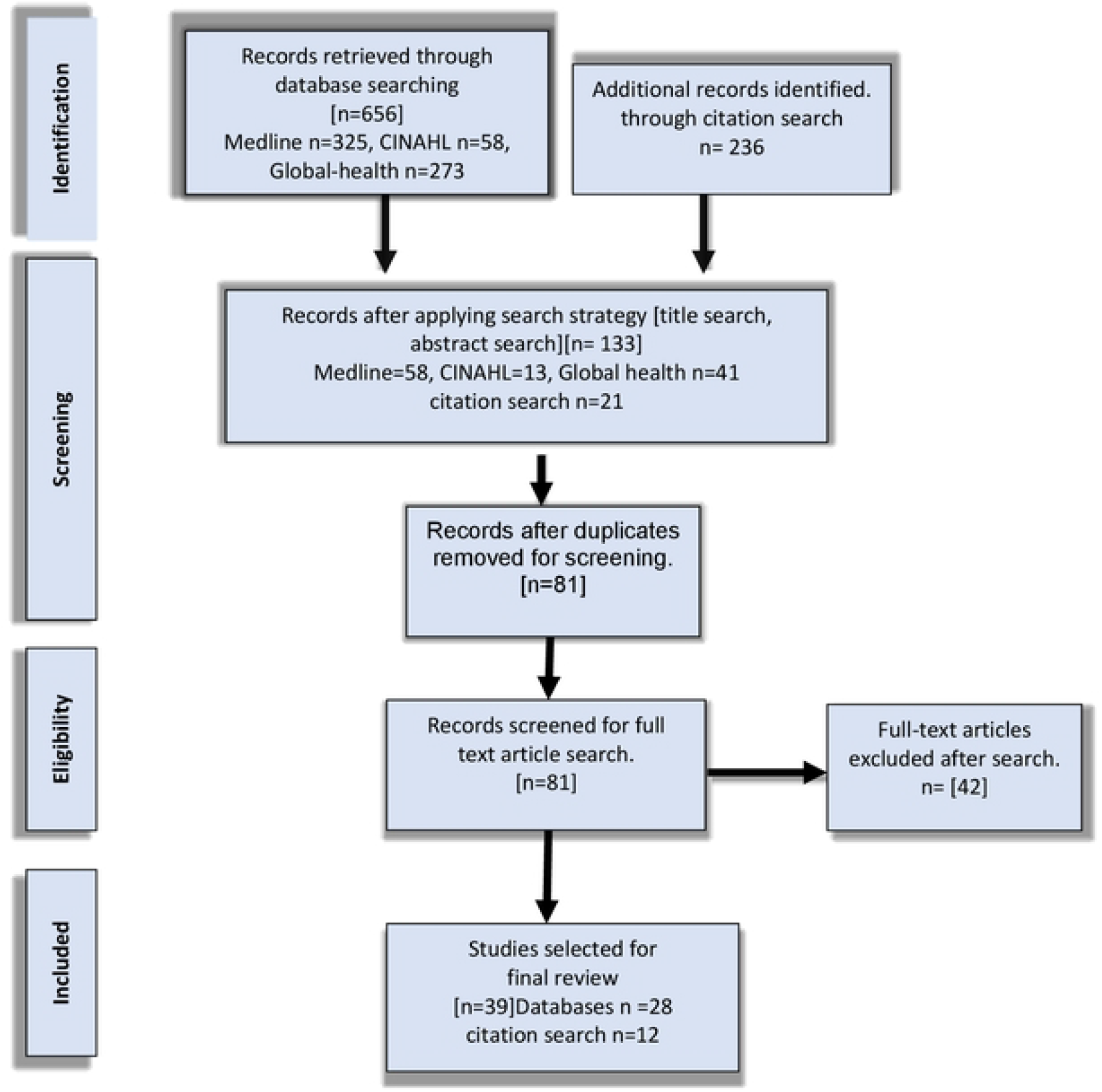
PRISMA flowchart showing the process of searching and selecting the relevant studies.

### Characteristics of included papers

Cumulatively, the 39 papers pertained to 19 MEC, with most [n=34] studies conducted in Africa [Table 2]. The remaining five studies were from Southeast Asia. Most publications (19/39) were led by authors affiliated with institutions in MEC, including nineteen from African institutions.

**Table 2:** Geographical distribution of selected studies.

| Region | Country | No. of studies | Total No. of studies |
| --- | --- | --- | --- |
| Asia | Bangladesh | 1 | 5 |
|  | Myanmar | 2 |  |
|  | Papua New Guinea | 1 |  |
|  | Thailand | 1 |  |
| Central Africa | DRC | 1 | 1 |
| East Africa | Ethiopia | 6 | 18 |
|  | Kenya | 1 |  |
|  | Madagascar | 1 |  |
|  | Rwanda | 1 |  |
|  | Tanzania | 4 |  |
|  | Uganda | 5 |  |
| West Africa | Benin | 1 | 11 |
|  | Burkina Faso | 1 |  |
|  | Ghana | 4 |  |
|  | Nigeria | 4 |  |
|  | Senegal | 1 |  |
| South Africa | Malawi | 1 | 2 |
|  | Zimbabwe | 1 |  |
| Multi-country | Ghana/Malawi/Kenya | 1 | 2 |
|  | Mali/Kenya | 1 |  |

**Table 3:**
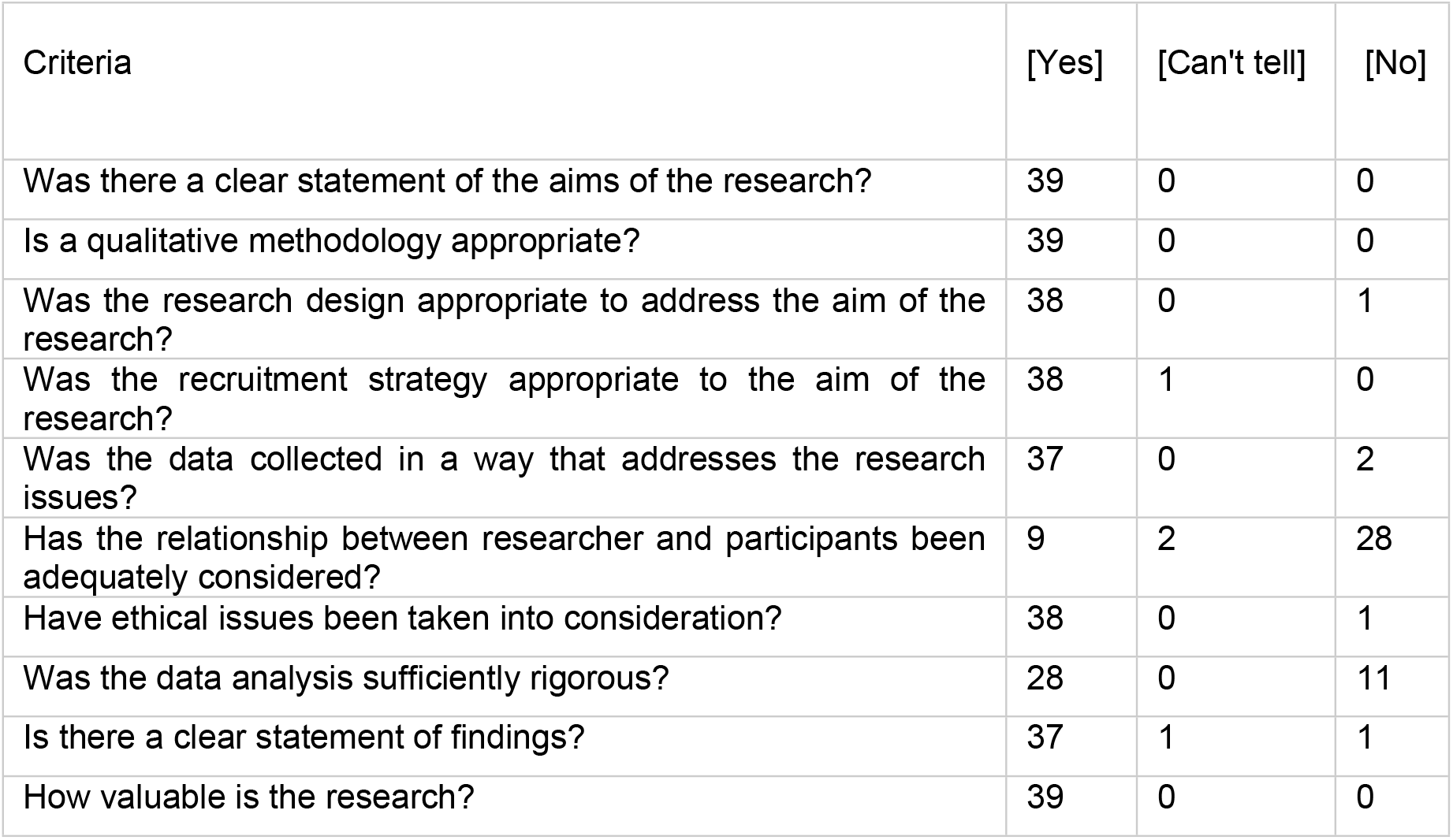
Quality assessment results.

| Criteria | [Yes] | [Can't tell] | [No] |
| --- | --- | --- | --- |
| Was there a clear statement of the aims of the research? | 39 | 0 | 0 |
| Is a qualitative methodology appropriate? | 39 | 0 | 0 |
| Was the research design appropriate to address the aim of the research? | 38 | 0 | 1 |
| Was the recruitment strategy appropriate to the aim of the research? | 38 | 1 | 0 |
| Was the data collected in a way that addresses the research issues? | 37 | 0 | 2 |
| Has the relationship between researcher and participants been adequately considered? | 9 | 2 | 28 |
| Have ethical issues been taken into consideration? | 38 | 0 | 1 |
| Was the data analysis sufficiently rigorous? | 28 | 0 | 11 |
| Is there a clear statement of findings? | 37 | 1 | 1 |
| How valuable is the research? | 39 | 0 | 0 |

A full description of the study characteristics of publications included in this review can be found in supplementary file one. As shown, over two-thirds [34] of the study populations were drawn from rural settings. Most studies reviewed [25/39] employed a qualitative study design alone, compared to a third [14/39] that used a mixed methods study design. Data collection methods variously included focus group discussions, key informant interviews, in-depth interviews, observation, and participatory activities. Sampling methods included snowballing, and convenient selection, although the majority used purposive sampling [20/39] to recruit study participants. More than two-thirds of studies [30] utilised thematic analysis.

### Quality assessment

Table 3 provides a summary of the quality assessment outcome. A CASP score of at least eight, considered to be high, was present in about 90% [35/39] of reviewed studies. The remaining 10% [4/39] received a score of six and seven, considered medium quality. No studies scored below five, and no publications were disregarded for this review based on the CASP score. As shown in Table 3, all studies [39/39] made clear their aims and objectives, and the majority clearly stated the suitability of their choice of a qualitative study design [38/39]. The recruitment strategy and data collection methods were appropriate in almost all reviewed papers [38/39 and 37/39, respectively]. Regarding reflexivity, nine studies provided details on how they evaluated their role, potential bias, influence during data collection, and how they reacted to events and any changes during the study. Even though 28 of the studies did not provide information on reflexivity, they did provide information on how they established relationships with study participants before the start of each survey. Almost all studies [38/39] indicated how ethical approval was obtained. In addition, most of the studies’ data analyses [28/39] were sufficiently thorough, and their findings [37/39] were clear and concise.

### Thematic analysis of key findings

Reported reasons for net non-use across the 39 publications included in the review were subsequently categorised under four main themes, each with two or more sub-themes. Each theme and sub-theme is presented in turn below, alongside relevant quotes taken from reviewed publications to illustrate key points. Supplementary file 2 lists the respective publications coded against each sub-theme.

#### Human factors

This theme refers to reasons for non-use that pertain to human perceptions, preferences or experiences. More than two-thirds of study findings were categorised under this theme, across the following six sub-themes:

#### Perceived effectiveness

Results suggest that participants do not always consider ITNs to be effective or considered other preventive methods to be more effective. For example, the absence of dead insects at the side of the mosquito nets gave some participants the impression that treated nets had become ineffective (17-21). Net cleanliness was also a factor in some cases(22). Fourteen studies had findings categorised under this sub-theme (23-29).

*“This net [an ITN] is more or less similar to an ordinary mosquito net because we found a mosquito with it even after hanging before sleeping” (17).*

*“If the area is fumigated even once a month by the government, then we will be free day and night from mosquito bite and not only in night” (28).*

*“Because of cleanliness, some husbands do not like dirty bed nets. They want nets to be washed every few days. So, if he gets home and finds that the bed net has not been washed, he will put it away. This may result in the family or couples sleeping without a bed net” (22).*

#### Discomfort

In nine studies, discomfort was presented as a reason for non-use, including the feeling of suffocation while sleeping under the nets, especially in a hot climate, or the fear of experiencing some reaction, such as skin irritation (26, 28, 30-35).

*“There’s many insecticides. Sometimes it’s too strong and difficult to sleep under the net for the first time. The insecticide prevents me from breathing well and it’s very difficult to breathe around this product” (34).*

*“I heard that one man slept inside the net and vomited blood,” and I heard that it causes skin irritation because the chemical is too much” (32).*

*“I am still afraid of malaria, but I could not sleep. It was too hot to sleep under the nets in the summer” (31).*

##### Seasonality of Mosquitoes

Reported in three studies, many participants felt that using nets during seasons when mosquitoes were less frequent, such as hot seasons, was unnecessary. At the same time, net use was more prevalent during rainy seasons when mosquitoes were perceived to be predominant (30, 34, 36):

*“We were given these nets during the rainy season when there were a lot of mosquitoes. I hung the nets, and the children slept in them that time. When the rainy season was over, we removed the nets because during that time, there was no mosquitoes” (36).*

##### Prioritising nets

In four studies, certain family members, such as infants, children under five, and pregnant women, were given priority access to available nets, with youth and older adults generally receiving less attention and prioritisation. The reason for prioritising mosquito nets could be associated with a lack of insecticide-treated nets in homes (22, 32, 37). In one study, the head of the house was prioritized over other family members(38).

*“I would not give priority to the youth because they are strong and their bodies can resist malaria… I would only consider those that are more vulnerable and leave those that are strong enough to fight” (38).*

*“We had in mind that the man, as the head of the family, should be the one to get it first; [he] has to sleep on the bed. If God provides more, then I and the kids shall get later” (38).*

#### Alternate net use

In six studies, alternate uses of mosquito nets such as covering farm animals, farm produce, processing milk from butter, catching fish, or rearing chickens were reported as a reason for not sleeping in them (19, 23, 35, 39-41)

*“The one I had following the distribution, frankly speaking, I didn’t sleep in it because it was hard. So, I used it for my windows. I fixed them at the back of my windows and even my trap door. That is what I used. I have used it as a net for all my windows so that mosquitoes do not enter my room” (35).*

*“We are in fear that what will happen in the future if we tell you everything… We use bed nets to cover the toilet, separating seeds from the stem; after thoroughly washing, we use them for filtering kinetic, coffee and milk during the separation process of milk from the butter. Those who cannot purchase clothes can use them as night clothes, as bed sheets, and it gives many more purposes” (19).*

##### Socio-cultural beliefs and practices

Social norms and practices can negatively influence the use of mosquito nets as highlighted in four studies. Outdoor nocturnal activities can make net use inpractical and, in two studies, individuals at a funeral were reportedly not allowed a mosquito net to sleep in as it was considered taboo or seen as disrespectful to the deceased and family (14, 25, 42, 43):

*“We don’t sleep under the net when it’s burial time… You cannot decide to put your net, who are you? How important are you? How arrogant are you? So, most of us in Teso don’t even sleep when at a funeral. We sit out around the fire or even within a house, and in big numbers, so one cannot use a mosquito net” (25).*

#### Net factors

This theme refers to reasons for non-use that relate to the physical properties of mosquito nets. Findings from 11 studies pertained to net factors, across two sub-themes.

##### Net preference

Seven studies reported that participants preference for certain characteristics of nets such as color, texture and size, infleunces net use (22, 26, 32, 35, 37, 40, 44). In two other studies, participants expressed how some features of the nets made them better suited for other domestic use, such as hanging over doors and windows or covering of livestock and farm produce (26, 35):

*“I have not yet installed the new net; it is still stored in its packaging with clothing. I do not like it because it has large meshes; I still use the old ones because they are smaller. In addition, the product on the new net gives the cold. The fabric is stiff and uncomfortable, my hair hangs in there”(37).*

*“I got a carpenter to fix the ITN over my bed, but he said it was too big and advised that I use it on my doors and windows. So he cut and fixed the net on my doors and windows. I also see people [using] the ITN as [a] fence around their farms and gardens to protect from chickens and some insects” (26).*

#### Net setup

In four studies, participants had difficulties with the initial or ongoing setup of the mosquito nets, which impacted on the use, including difficulty in getting into the nets (20, 27, 31, 35):

*“Once they fix the net and find entering the net uncomfortable, they will not sleep in it again because it did not serve their purpose. In the night, if they want to go and urinate and the net ties them up and they have to remove it, go out and come back to fix it, they feel like they are in prison and will not sleep under it*.*”(35).*

#### Environmental/lifestyle factors

This theme refers to factors in the immediate environment or participants’ lifestyle that contributed towards net non-use among study participants.Findings from seven studies pertained to environmental/lifestyle factors, across two sub-themes.

#### Travel or nocturnal activity

As reported in five studies, mosquito net use away from one’s primary residence can be challenging, particularly for individuals active at night or who frequently travel for businesses or social engagements. As a result, participants in such situations find it difficult to use bed nets and are often at risk for malaria (14, 22, 25, 40, 42):

*“Sometimes when we go farming in the valleys [protection from mosquitoes is not possible]. You might go with the intention of coming back but find that it gets dark, and so you decide to sleep there”(42).*

*“Sometimes we travel, and some homes that host us may have no bed nets, so when you stay for some days in these home,s you are likely to get malaria”(22).*

#### Architecture of homes

In two studies, the structures and style of homes/houses played an important role in determining participants continuos use of mosquito nets. Participants living in houses built on stilts, for example were unable to use mosquito nets properly due to the gaps in the floorboards that allow mosquitoes to enter, regardless of the number of nets distributed to them (27, 45):

*“Because our houses are built with wood on stilts, there is space between the planks. Even if the bed net is well tied to the ceiling, there is always a space beneath the planks that mosquito harnesses to get in. But I don’t think there is a way to avoid mosquitoes unless we change our houses. That would require a lot of money. The best way to help us is to change our houses” (42).*

#### Administrative/Economic Factors

This theme refers to a participant’s ability to access or purchase mosquito nets. Findings from nine studies pertained to administaryive/economic factors, across two sub-themes.

##### Accessing nets

Five studies highlited participants dependence on free net distribution, especially in health facilties, with reports of bias in such distributions resulting in adequate numbers of nets available within the household (11, 17, 24, 46, 47).

*“It is segregation, favouritism, and all kind of things. For example, if they received 40 Bednets, doctors give them to 5-6 people who did not visit the health centre. If we get there, they said that is it finished” (48).*

*“These days, people get bed nets at the health centre from antenatal consultation or child vaccination at nine months. However, those who don’t attend consultation or vaccination services have a problem with getting bed nets” (24).*

#### Economic implications

Participants in four studies who could not obtain nets via free or subsidised distribution campaigns either had to raise money to procure new nets, or stay without them (22, 36, 45, 49).

*“ITNs are sold at the health facility at a lower price. If you don’t go early, you might not get [sic] it to buy because a lot of people go there to get them. If you miss this, then you have to buy it from the open market at a relatively higher price. So, if you don’t have the money, you cannot get the net to use” (49).*

*Nets received via free distribution campaigns were exchanged for money in some cases. One reason given for such gestures was poverty (36, 45)*.

*“We ensure free bed net distribution to pregnant women and young children, but the issue is that many of these women sell their nets” (45).*

## DISCUSSION

In this review, we sought to assess the extent and quality of qualitative research published between 2011 and 2021 pertaining to the non-use of all types of mosquito nets and, drawing on the findings from this literature, update current understanding of reasons for mosquito net non-use in MEC. The focus on qualitative research was informed by an earlier review published in 2011 which identified a dearth of qualitative investigation on this topic. Our review identified substantial growth in qualitative research on the topic of mosquito net non-use with 39 qualitative or mixed methods studies published between 2011 and 2021 as compared to four between 1999 and 2010 (12). Significantly, this growth in qualitative research was primarily driven by researchers working at institutions located in MEC.

Not only did we find growth in published, qualitative research output, but our findings also indicate the increased research output was of a generally high standard. A CASP score of at least eight out of ten, indicative of high quality, was awarded to more than 90% of the reviewed publications. As MEC-based researchers primarily drove the growth in qualitative research publication, this finding is especially pleasing concerning research capacity and equity issues in global health research (50). Nevertheless, two-thirds of the reviewed papers [28/39] were rated poorly on the CASP measure of reflexivity [Table 4], suggesting this may be an aspect that needs greater attention when preparing qualitative research for publication, potentially through effective strategies such as reflexive writing or collaborative reflection (51).

**Table 4:** Summary of emerging themes.

| Analytical/<br>final theme | Sub-theme | Descriptive theme |
| --- | --- | --- |
| Human factor | Perceived effectiveness | In-effectiveness of nets<br>Attitudes to net use<br>Preference for other preventive methods<br>Net hygiene |
|  | Discomfort | Adverse effect of insecticides<br>Hot climate |
|  | Seasonality of Mosquito | Perceived low density of mosquito |
|  | Prioritizing nets | Sleeping arrangement |
|  | Alternate net use | Domestic use of net |
|  | Socio-cultural practices | Social gatherings |
| Net factor | Net preference | Characteristics of nets<br>Improper use of net |
|  | Net set up | Challenges with Net placement |
| Environmental/<br>lifestyle factor | Architecture of home | Home structure |
|  | Travel or nocturnal activities | Net use away from home |
| Administrative/<br>economic<br>factor | Access to nets | Net distribution |
|  | Economic implications | New net purchase<br>Net exchange for money |

With regards to updating current understanding of reasons for mosquito net non-use in MEC, our study re-affirms many of the findings presented in the original review by Pulford et al. (12). The two most frequently cited reasons for mosquito net non-use determined by Pulford et al: discomfort due to heat and perceived low mosquito density were also prominent in many of the qualitative studies included in our review. The continued reporting of these factors over an extended period via quantitative, and now a more significant number of qualitative studies, strongly suggests that they are persistent and common barriers to mosquito net use. However, our findings also extend upon those presented in the original Pulford et al. review, primarily through the identification of four overarching factors that contribute towards net non-use; namely, human factors, net factors, environmental/lifestyle factors and administrative/economic factors. The human and the net factor were the two most commonly reported factors. Interventions that might address these issues include continuous education aimed at changing human behaviour and interventions aimed at designing mosquito nets that are less harmful and more user-friendly in terms of style, size, and texture.

The present study is the first literature review that exclusively focuses on qualitative research on the reported reasons for the non-use of all forms of mosquito nets. The included papers are proven to be of high quality. Two independent researchers validated the article screening, final theme selection, and quality assessment, which we believe reduced selection bias. The synthesis findings were supported by multiple quotes from the various studies, representing a complete description of the themes, which increased trustworthiness. Despite these efforts to ensure quality, limitations remained. Only English language publications were included, and grey literature was excluded. Thus, we may have left out qualitative data related to the causes of mosquito net non-use that could have increased the value of our research results. The similarity in geographical distribution and economic status of the MEC in the review enhances the transferability of findings. However, the debate over decontextualizing qualitative synthesis methods may limit the transferability of the results (52).A further review of quantitative research may be required to generalise the findings.

## Data Availability

Data included in manuscript.

## Conclusion

Over the last decade, there has been a significant increase in high-quality qualitative research, contributing to a consolidated and more in-depth understanding of the reasons for mosquito net non-use. The review findings highlight the wide range of factors that influence net use. Yet, some factors have been consistently reported at high frequency over an extended time period indicating these are priority concerns to address. The research focus should shift toward intervention studies to address these issues.

## Acknowledgments

The Mamco Selab scholarship at the Liverpool School of Tropical Medicine for part tuition fee in support of a Masters’s degree programme for which this literature review was completed.

## Contributors

HL, and JP were involved in designing the review. HL was the lead researcher, actively engaged in searching, screening, quality assessment, and data analysis. JP conceptualised the study. US participated in the screening and quality assessment of the relevant papers.

## Patient consent for publication

Not required

## Data sharing statement

All data will be made available upon request

## Supporting information

S5 Table: Summary of study’s characteristics.

S6 Table: Lead Authors Instituiton of affliation.

S7 Table: Characteristics of included studies.

S8 Table: Result of synthesis.

